## Supplementary file 1 for "Nuclear magnetic resonance-based metabolomics with machine learning for predicting progression from prediabetes to diabetes"

**Supplementary file 1. List of 168 NMR-based metabolomic biomarkers in the UK Biobank.**

| **Field ID** | **Metabolites** | **Unit** | **Group** |
| --- | --- | --- | --- |
| 23427 | Total Lipoprotein Particles | mmol/l | Lipoprotein particle |
| 23428 | Total VLDL Particles | mmol/l | Lipoprotein particle |
| 23481 | Chylomicrons and Extremely Large VLDL Particles | mmol/l | Lipoprotein particle |
| 23488 | Very Large VLDL Particles | mmol/l | Lipoprotein particle |
| 23495 | Large VLDL Particles | mmol/l | Lipoprotein particle |
| 23502 | Medium VLDL Particles | mmol/l | Lipoprotein particle |
| 23509 | Small VLDL Particles | mmol/l | Lipoprotein particle |
| 23516 | Very Small VLDL Particles | mmol/l | Lipoprotein particle |
| 23429 | Total LDL Particles | mmol/l | Lipoprotein particle |
| 23523 | IDL Particles | mmol/l | Lipoprotein particle |
| 23530 | Large LDL Particles | mmol/l | Lipoprotein particle |
| 23537 | Medium LDL Particles | mmol/l | Lipoprotein particle |
| 23544 | Small LDL Particles | mmol/l | Lipoprotein particle |
| 23430 | Total HDL Particles | mmol/l | Lipoprotein particle |
| 23551 | Very Large HDL Particles | mmol/l | Lipoprotein particle |
| 23558 | Large HDL Particles | mmol/l | Lipoprotein particle |
| 23565 | Medium HDL Particles | mmol/l | Lipoprotein particle |
| 23572 | Small HDL Particles | mmol/l | Lipoprotein particle |
| 23400 | Total Cholesterol | mmol/l | Cholesterol |
| 23401 | None-HDL Cholesterol (Total Cholesterol Minus HDL-C) | mmol/l | Cholesterol |
| 23402 | Remnant Cholesterol (Non-HDL, Non-LDL -Cholesterol) | mmol/l | Cholesterol |
| 23403 | Total VLDL Cholesterol | mmol/l | Cholesterol |
| 23484 | Cholesterol in Chylomicrons and Extremely Large VLDL | mmol/l | Cholesterol |
| 23491 | Cholesterol in Very Large VLDL | mmol/l | Cholesterol |
| 23498 | Cholesterol in Large VLDL | mmol/l | Cholesterol |
| 23505 | Cholesterol in Medium VLDL | mmol/l | Cholesterol |
| 23512 | Cholesterol in Small VLDL | mmol/l | Cholesterol |
| 23519 | Cholesterol in Very Small VLDL | mmol/l | Cholesterol |
| 23405 | Total LDL Cholesterol | mmol/l | Cholesterol |
| 23404 | Clinical LDL Cholesterol | mmol/l | Cholesterol |
| 23526 | Cholesterol in IDL | mmol/l | Cholesterol |
| 23533 | Cholesterol in Large LDL | mmol/l | Cholesterol |
| 23540 | Cholesterol in Medium LDL | mmol/l | Cholesterol |
| 23547 | Cholesterol in Small LDL | mmol/l | Cholesterol |
| 23406 | Total HDL Cholesterol | mmol/l | Cholesterol |
| 23554 | Cholesterol in Very Large HDL | mmol/l | Cholesterol |
| 23561 | Cholesterol in Large HDL | mmol/l | Cholesterol |
| 23568 | Cholesterol in Medium HDL | mmol/l | Cholesterol |
| 23575 | Cholesterol in Small HDL | mmol/l | Cholesterol |
| 23419 | Total Free Cholesterol | mmol/l | Free cholesterol |
| 23420 | Total Free Cholesterol in VLDL | mmol/l | Free cholesterol |
| 23486 | Free Cholesterol in Chylomicrons and Extremely Large VLDL | mmol/l | Free cholesterol |
| 23493 | Free Cholesterol in Very Large VLDL | mmol/l | Free cholesterol |
| 23500 | Free Cholesterol in Large VLDL | mmol/l | Free cholesterol |
| 23507 | Free Cholesterol in Medium VLDL | mmol/l | Free cholesterol |
| 23514 | Free Cholesterol in Small VLDL | mmol/l | Free cholesterol |
| 23521 | Free Cholesterol in Very Small VLDL | mmol/l | Free cholesterol |
| 23421 | Total Free Cholesterol in LDL | mmol/l | Free cholesterol |
| 23528 | Free Cholesterol in IDL | mmol/l | Free cholesterol |
| 23535 | Free Cholesterol in Large LDL | mmol/l | Free cholesterol |
| 23542 | Free Cholesterol in Medium LDL | mmol/l | Free cholesterol |
| 23549 | Free Cholesterol in Small LDL | mmol/l | Free cholesterol |
| 23422 | Total Free Cholesterol in HDL | mmol/l | Free cholesterol |
| 23556 | Free Cholesterol in Very Large HDL | mmol/l | Free cholesterol |
| 23563 | Free Cholesterol in Large HDL | mmol/l | Free cholesterol |
| 23570 | Free Cholesterol in Medium HDL | mmol/l | Free cholesterol |
| 23577 | Free Cholesterol in Small HDL | mmol/l | Free cholesterol |
| 23415 | Total Esterified Cholesterol | mmol/l | Cholesteryl Esters |
| 23416 | Total Cholesteryl Esters in VLDL | mmol/l | Cholesteryl Esters |
| 23485 | Cholesteryl Esters in Chylomicrons and Extremely Large VLDL | mmol/l | Cholesteryl Esters |
| 23492 | Cholesteryl Esters in Very Large VLDL | mmol/l | Cholesteryl Esters |
| 23499 | Cholesteryl Esters in Large VLDL | mmol/l | Cholesteryl Esters |
| 23506 | Cholesteryl Esters in Medium VLDL | mmol/l | Cholesteryl Esters |
| 23513 | Cholesteryl Esters in Small VLDL | mmol/l | Cholesteryl Esters |
| 23520 | Cholesteryl Esters in Very Small VLDL | mmol/l | Cholesteryl Esters |
| 23417 | Total Cholesteryl Esters in LDL | mmol/l | Cholesteryl Esters |
| 23527 | Cholesteryl Esters in IDL | mmol/l | Cholesteryl Esters |
| 23534 | Cholesteryl Esters in Large LDL | mmol/l | Cholesteryl Esters |
| 23541 | Cholesteryl Esters in Medium LDL | mmol/l | Cholesteryl Esters |
| 23548 | Cholesteryl Esters in Small LDL | mmol/l | Cholesteryl Esters |
| 23418 | Total Cholesteryl Esters in HDL | mmol/l | Cholesteryl Esters |
| 23555 | Cholesteryl Esters in Very Large HDL | mmol/l | Cholesteryl Esters |
| 23562 | Cholesteryl Esters in Large HDL | mmol/l | Cholesteryl Esters |
| 23569 | Cholesteryl Esters in Medium HDL | mmol/l | Cholesteryl Esters |
| 23576 | Cholesteryl Esters in Small HDL | mmol/l | Cholesteryl Esters |
| 23407 | Total Triglycerides | mmol/l | Triglycerides |
| 23408 | Total Triglycerides in VLDL | mmol/l | Triglycerides |
| 23487 | Triglycerides in Chylomicrons and Extremely Large VLDL | mmol/l | Triglycerides |
| 23494 | Triglycerides in Very Large VLDL | mmol/l | Triglycerides |
| 23501 | Triglycerides in Large VLDL | mmol/l | Triglycerides |
| 23508 | Triglycerides in Medium VLDL | mmol/l | Triglycerides |
| 23515 | Triglycerides in Small VLDL | mmol/l | Triglycerides |
| 23522 | Triglycerides in Very Small VLDL | mmol/l | Triglycerides |
| 23409 | Total Triglycerides in LDL | mmol/l | Triglycerides |
| 23529 | Triglycerides in IDL | mmol/l | Triglycerides |
| 23536 | Triglycerides in Large LDL | mmol/l | Triglycerides |
| 23543 | Triglycerides in Medium LDL | mmol/l | Triglycerides |
| 23550 | Triglycerides in Small LDL | mmol/l | Triglycerides |
| 23410 | Total Triglycerides in HDL | mmol/l | Triglycerides |
| 23557 | Triglycerides in Very Large HDL | mmol/l | Triglycerides |
| 23564 | Triglycerides in Large HDL | mmol/l | Triglycerides |
| 23571 | Triglycerides in Medium HDL | mmol/l | Triglycerides |
| 23578 | Triglycerides in Small HDL | mmol/l | Triglycerides |
| 23411 | Total Phospholipids in Lipoprotein Particles | mmol/l | Phospholipids |
| 23412 | Total Phospholipids in VLDL | mmol/l | Phospholipids |
| 23483 | Phospholipids in Chylomicrons and Extremely Large VLDL | mmol/l | Phospholipids |
| 23490 | Phospholipids in Very Large VLDL | mmol/l | Phospholipids |
| 23497 | Phospholipids in Large VLDL | mmol/l | Phospholipids |
| 23504 | Phospholipids in Medium VLDL | mmol/l | Phospholipids |
| 23511 | Phospholipids in Small VLDL | mmol/l | Phospholipids |
| 23518 | Phospholipids in Very Small VLDL | mmol/l | Phospholipids |
| 23413 | Total Phospholipids in LDL | mmol/l | Phospholipids |
| 23525 | Phospholipids in IDL | mmol/l | Phospholipids |
| 23532 | Phospholipids in Large LDL | mmol/l | Phospholipids |
| 23539 | Phospholipids in Medium LDL | mmol/l | Phospholipids |
| 23546 | Phospholipids in Small LDL | mmol/l | Phospholipids |
| 23414 | Total Phospholipids in HDL | mmol/l | Phospholipids |
| 23553 | Phospholipids in Very Large HDL | mmol/l | Phospholipids |
| 23560 | Phospholipids in Large HDL | mmol/l | Phospholipids |
| 23567 | Phospholipids in Medium HDL | mmol/l | Phospholipids |
| 23574 | Phospholipids in Small HDL | mmol/l | Phospholipids |
| 23423 | Total Lipids in Lipoprotein Particles | mmol/l | Total Lipids |
| 23424 | Total Lipids in VLDL | mmol/l | Total Lipids |
| 23482 | Total Lipids in Chylomicrons and Extremely Large VLDL | mmol/l | Total Lipids |
| 23489 | Total Lipids in Very Large VLDL | mmol/l | Total Lipids |
| 23496 | Total Lipids in Large VLDL | mmol/l | Total Lipids |
| 23503 | Total Lipids in Medium VLDL | mmol/l | Total Lipids |
| 23510 | Total Lipids in Small VLDL | mmol/l | Total Lipids |
| 23517 | Total Lipids in Very Small VLDL | mmol/l | Total Lipids |
| 23425 | Total Lipids in LDL | mmol/l | Total Lipids |
| 23524 | Total Lipids in IDL | mmol/l | Total Lipids |
| 23531 | Total Lipids in Large LDL | mmol/l | Total Lipids |
| 23538 | Total Lipids in Medium LDL | mmol/l | Total Lipids |
| 23545 | Total Lipids in Small LDL | mmol/l | Total Lipids |
| 23426 | Total Lipids in HDL | mmol/l | Total Lipids |
| 23552 | Total Lipids in Very Large HDL | mmol/l | Total Lipids |
| 23559 | Total Lipids in Large HDL | mmol/l | Total Lipids |
| 23566 | Total Lipids in Medium HDL | mmol/l | Total Lipids |
| 23573 | Total Lipids in Small HDL | mmol/l | Total Lipids |
| 23431 | Average Diameter for VLDL Particles | nm | Sizes & apolipoprotein |
| 23432 | Average Diameter for LDL Particles | nm | Sizes & apolipoprotein |
| 23433 | Average Diameter for HDL Particles | nm | Sizes & apolipoprotein |
| 23440 | Apolipoprotein A1 | g/l | Sizes & apolipoprotein |
| 23439 | Apolipoprotein B | g/l | Sizes & apolipoprotein |
| 23442 | Total Fatty Acids | mmol/l | Fatty acids |
| 23444 | Omega-3 Fatty Acids | mmol/l | Fatty acids |
| 23445 | Omega-6 Fatty Acids | mmol/l | Fatty acids |
| 23446 | Polyunsaturated Fatty Acids | mmol/l | Fatty acids |
| 23447 | Monounsaturated Fatty Acids | mmol/l | Fatty acids |
| 23448 | Saturated Fatty Acids | mmol/l | Fatty acids |
| 23449 | Linoleic Acid | mmol/l | Fatty acids |
| 23450 | Docosahexaenoic Acid | mmol/l | Fatty acids |
| 23443 | Degree of Unsaturation |  | Fatty acids |
| 23436 | Total Cholines | mmol/l | Amino acids, cholines, and glycolysis |
| 23437 | Phosphatidylcholines | mmol/l | Amino acids, cholines, and glycolysis |
| 23438 | Sphingomyelins | mmol/l | Amino acids, cholines, and glycolysis |
| 23434 | Phosphoglycerides | mmol/l | Amino acids, cholines, and glycolysis |
| 23470 | Glucose | mmol/l | Amino acids, cholines, and glycolysis |
| 23471 | Lactate | mmol/l | Amino acids, cholines, and glycolysis |
| 23472 | Pyruvate | mmol/l | Amino acids, cholines, and glycolysis |
| 23473 | Citrate | mmol/l | Amino acids, cholines, and glycolysis |
| 23460 | Alanine | mmol/l | Amino acids, cholines, and glycolysis |
| 23461 | Glutamine | mmol/l | Amino acids, cholines, and glycolysis |
| 23462 | Glycine | mmol/l | Amino acids, cholines, and glycolysis |
| 23463 | Histidine | mmol/l | Amino acids, cholines, and glycolysis |
| 23465 | Isoleucine | mmol/l | Amino acids, cholines, and glycolysis |
| 23466 | Leucine | mmol/l | Amino acids, cholines, and glycolysis |
| 23467 | Valine | mmol/l | Amino acids, cholines, and glycolysis |
| 23468 | Phenylalanine | mmol/l | Amino acids, cholines, and glycolysis |
| 23469 | Tyrosine | mmol/l | Amino acids, cholines, and glycolysis |
| 23464 | Branched-Chain Amino Acids (Leucine + Isoleucine + Valine) | mmol/l | Amino acids, cholines, and glycolysis |
| 23474 | 3-Hydroxybutyrate | mmol/l | Ketone bodies & fluid balance |
| 23475 | Acetate | mmol/l | Ketone bodies & fluid balance |
| 23476 | Acetoacetate | mmol/l | Ketone bodies & fluid balance |
| 23477 | Acetone | mmol/l | Ketone bodies & fluid balance |
| 23479 | Albumin | g/l | Ketone bodies & fluid balance |
| 23478 | Creatinine | mmol/l | Ketone bodies & fluid balance |
| 23480 | Glycoprotein Acetyls | mmol/l | Ketone bodies & fluid balance |

HDL, high-density lipoproteins; IDL, intermediate-density lipoproteins; LDL, low-density lipoproteins; VLDL, very-low-density lipoproteins.
