## Supplementary file 2 for "Nuclear magnetic resonance-based metabolomics with machine learning for predicting progression from prediabetes to diabetes"

**Supplementary file 2. Associations of 168 metabolic biomarkers with risk of diabetes among 13,489 participants with prediabetes.**

| **Metabolites** | **HR (95% CI)** | ***P* value** |
| --- | --- | --- |
| **Lipoprotein particle** |  |  |
| Total Lipoprotein Particles | 0.94 (0.90, 0.98) | 5.50E-03 |
| Total VLDL Particles | 1.15 (1.11, 1.20) | **5.30E-12** |
| Chylomicrons and Extremely Large VLDL Particles | 1.24 (1.20, 1.29) | **1.23E-34** |
| Very Large VLDL Particles | 1.24 (1.20, 1.29) | **1.10E-30** |
| Large VLDL Particles | 1.22 (1.18, 1.27) | **3.62E-25** |
| Medium VLDL Particles | 1.09 (1.04, 1.13) | **1.22E-04** |
| Small VLDL Particles | 1.17 (1.12, 1.21) | **1.16E-14** |
| Very Small VLDL Particles | 1.09 (1.05, 1.14) | **3.30E-05** |
| Total LDL Particles | 1.00 (0.96, 1.05) | 9.42E-01 |
| IDL Particles | 0.97 (0.93, 1.01) | 1.76E-01 |
| Large LDL Particles | 0.98 (0.94, 1.02) | 3.08E-01 |
| Medium LDL Particles | 1.03 (0.99, 1.08) | 1.54E-01 |
| Small LDL Particles | 1.05 (1.01, 1.10) | 1.55E-02 |
| Total HDL Particles | 0.93 (0.89, 0.98) | 2.44E-03 |
| Very Large HDL Particles | 0.84 (0.79, 0.88) | **3.38E-10** |
| Large HDL Particles | 0.78 (0.74, 0.83) | **1.41E-16** |
| Medium HDL Particles | 0.94 (0.90, 0.99) | 1.14E-02 |
| Small HDL Particles | 1.00 (0.96, 1.04) | 8.80E-01 |
| **Cholesterol** |  |  |
| Total Cholesterol | 0.94 (0.90, 0.99) | 8.88E-03 |
| None-HDL Cholesterol (Total Cholesterol Minus HDL-C) | 0.99 (0.95, 1.03) | 6.20E-01 |
| Remnant Cholesterol (Non-HDL, Non-LDL -Cholesterol) | 1.02 (0.98, 1.06) | 4.07E-01 |
| Total VLDL Cholesterol | 1.11 (1.07, 1.16) | **6.35E-07** |
| Cholesterol in Chylomicrons and Extremely Large VLDL | 1.24 (1.20, 1.29) | **2.11E-29** |
| Cholesterol in Very Large VLDL | 1.21 (1.16, 1.26) | **8.51E-20** |
| Cholesterol in Large VLDL | 1.19 (1.15, 1.24) | **2.14E-17** |
| Cholesterol in Medium VLDL | 0.96 (0.92, 1.01) | 9.89E-02 |
| Cholesterol in Small VLDL | 1.07 (1.03, 1.12) | 1.27E-03 |
| Cholesterol in Very Small VLDL | 1.00 (0.96, 1.05) | 9.63E-01 |
| Total LDL Cholesterol | 0.96 (0.92, 1.00) | 7.96E-02 |
| Clinical LDL Cholesterol | 0.93 (0.89, 0.97) | 1.66E-03 |
| Cholesterol in IDL | 0.91 (0.87, 0.95) | **1.38E-05** |
| Cholesterol in Large LDL | 0.94 (0.90, 0.98) | 5.69E-03 |
| Cholesterol in Medium LDL | 1.00 (0.96, 1.05) | 8.62E-01 |
| Cholesterol in Small LDL | 1.00 (0.96, 1.04) | 9.00E-01 |
| Total HDL Cholesterol | 0.83 (0.79, 0.87) | **2.34E-14** |
| Cholesterol in Very Large HDL | 0.77 (0.73, 0.81) | **9.61E-20** |
| Cholesterol in Large HDL | 0.75 (0.71, 0.80) | **3.22E-23** |
| Cholesterol in Medium HDL | 0.89 (0.85, 0.94) | **1.15E-06** |
| Cholesterol in Small HDL | 0.98 (0.94, 1.02) | 2.41E-01 |
| **Free cholesterol** |  |  |
| Total Free Cholesterol | 0.98 (0.93, 1.02) | 2.74E-01 |
| Total Free Cholesterol in VLDL | 1.16 (1.11, 1.21) | **1.25E-12** |
| Free Cholesterol in Chylomicrons and Extremely Large VLDL | 1.25 (1.20, 1.29) | **2.03E-32** |
| Free Cholesterol in Very Large VLDL | 1.23 (1.19, 1.28) | **2.23E-26** |
| Free Cholesterol in Large VLDL | 1.22 (1.17, 1.27) | **2.76E-23** |
| Free Cholesterol in Medium VLDL | 1.04 (1.00, 1.09) | 5.29E-02 |
| Free Cholesterol in Small VLDL | 1.03 (0.99, 1.08) | 1.42E-01 |
| Free Cholesterol in Very Small VLDL | 1.06 (1.02, 1.11) | 4.64E-03 |
| Total Free Cholesterol in LDL | 0.90 (0.86, 0.94) | **3.50E-06** |
| Free Cholesterol in IDL | 0.89 (0.86, 0.94) | **1.51E-06** |
| Free Cholesterol in Large LDL | 0.89 (0.85, 0.93) | **5.86E-07** |
| Free Cholesterol in Medium LDL | 0.93 (0.89, 0.97) | 3.92E-04 |
| Free Cholesterol in Small LDL | 0.91 (0.87, 0.95) | **4.60E-06** |
| Total Free Cholesterol in HDL | 0.89 (0.85, 0.94) | **3.88E-06** |
| Free Cholesterol in Very Large HDL | 0.84 (0.80, 0.89) | **7.64E-11** |
| Free Cholesterol in Large HDL | 0.80 (0.76, 0.85) | **9.66E-15** |
| Free Cholesterol in Medium HDL | 0.93 (0.89, 0.98) | 2.53E-03 |
| Free Cholesterol in Small HDL | 1.05 (1.01, 1.09) | 1.69E-02 |
| **Cholesteryl Esters** |  |  |
| Total Esterified Cholesterol | 0.93 (0.89, 0.97) | 1.33E-03 |
| Total Cholesteryl Esters in VLDL | 1.07 (1.03, 1.12) | 1.40E-03 |
| Cholesteryl Esters in Chylomicrons and Extremely Large VLDL | 1.23 (1.18, 1.28) | **1.34E-26** |
| Cholesteryl Esters in Very Large VLDL | 1.16 (1.12, 1.21) | **5.10E-13** |
| Cholesteryl Esters in Large VLDL | 1.15 (1.10, 1.20) | **3.15E-11** |
| Cholesteryl Esters in Medium VLDL | 0.90 (0.86, 0.95) | **1.28E-05** |
| Cholesteryl Esters in Small VLDL | 1.09 (1.05, 1.14) | **2.83E-05** |
| Cholesteryl Esters in Very Small VLDL | 0.97 (0.93, 1.02) | 2.47E-01 |
| Total Cholesteryl Esters in LDL | 0.99 (0.95, 1.03) | 5.22E-01 |
| Cholesteryl Esters in IDL | 0.91 (0.87, 0.95) | **3.51E-05** |
| Cholesteryl Esters in Large LDL | 0.96 (0.92, 1.00) | 5.75E-02 |
| Cholesteryl Esters in Medium LDL | 1.03 (0.99, 1.08) | 1.21E-01 |
| Cholesteryl Esters in Small LDL | 1.04 (0.99, 1.08) | 1.10E-01 |
| Total Cholesteryl Esters in HDL | 0.81 (0.77, 0.85) | **3.67E-17** |
| Cholesteryl Esters in Very Large HDL | 0.75 (0.71, 0.80) | **5.66E-22** |
| Cholesteryl Esters in Large HDL | 0.74 (0.70, 0.78) | **1.23E-25** |
| Cholesteryl Esters in Medium HDL | 0.88 (0.85, 0.93) | **1.12E-07** |
| Cholesteryl Esters in Small HDL | 0.95 (0.91, 0.99) | 1.80E-02 |
| **Triglycerides** |  |  |
| Total Triglycerides | 1.26 (1.21, 1.31) | **1.23E-29** |
| Total Triglycerides in VLDL | 1.25 (1.20, 1.30) | **3.30E-28** |
| Triglycerides in Chylomicrons and Extremely Large VLDL | 1.25 (1.21, 1.30) | **1.76E-32** |
| Triglycerides in Very Large VLDL | 1.21 (1.17, 1.26) | **8.65E-23** |
| Triglycerides in Large VLDL | 1.20 (1.15, 1.25) | **1.85E-19** |
| Triglycerides in Medium VLDL | 1.24 (1.19, 1.28) | **5.09E-28** |
| Triglycerides in Small VLDL | 1.23 (1.18, 1.27) | **5.13E-30** |
| Triglycerides in Very Small VLDL | 1.22 (1.18, 1.26) | **3.83E-27** |
| Total Triglycerides in LDL | 1.21 (1.17, 1.26) | **1.26E-25** |
| Triglycerides in IDL | 1.21 (1.16, 1.25) | **7.24E-25** |
| Triglycerides in Large LDL | 1.22 (1.18, 1.26) | **1.43E-28** |
| Triglycerides in Medium LDL | 1.23 (1.19, 1.27) | **1.99E-31** |
| Triglycerides in Small LDL | 1.24 (1.19, 1.28) | **4.13E-31** |
| Total Triglycerides in HDL | 1.18 (1.14, 1.22) | **5.92E-22** |
| Triglycerides in Very Large HDL | 1.16 (1.12, 1.20) | **3.69E-15** |
| Triglycerides in Large HDL | 1.24 (1.19, 1.28) | **1.98E-30** |
| Triglycerides in Medium HDL | 1.28 (1.24, 1.33) | **1.26E-39** |
| Triglycerides in Small HDL | 1.21 (1.16, 1.25) | **7.24E-25** |
| **Phospholipids** |  |  |
| Total Phospholipids in Lipoprotein Particles | 1.03 (0.99, 1.08) | 1.45E-01 |
| Total Phospholipids in VLDL | 1.19 (1.15, 1.24) | **2.99E-17** |
| Phospholipids in Chylomicrons and Extremely Large VLDL | 1.25 (1.21, 1.30) | **4.74E-34** |
| Phospholipids in Very Large VLDL | 1.24 (1.19, 1.29) | **8.46E-29** |
| Phospholipids in Large VLDL | 1.24 (1.19, 1.29) | **1.80E-26** |
| Phospholipids in Medium VLDL | 1.08 (1.04, 1.13) | **2.43E-04** |
| Phospholipids in Small VLDL | 1.09 (1.05, 1.14) | **3.35E-05** |
| Phospholipids in Very Small VLDL | 1.12 (1.07, 1.16) | **1.24E-07** |
| Total Phospholipids in LDL | 0.97 (0.93, 1.01) | 1.61E-01 |
| Phospholipids in IDL | 0.95 (0.91, 0.99) | 2.18E-02 |
| Phospholipids in Large LDL | 0.94 (0.90, 0.99) | 9.52E-03 |
| Phospholipids in Medium LDL | 1.01 (0.97, 1.05) | 6.81E-01 |
| Phospholipids in Small LDL | 1.00 (0.96, 1.04) | 9.42E-01 |
| Total Phospholipids in HDL | 0.96 (0.92, 1.00) | 6.98E-02 |
| Phospholipids in Very Large HDL | 0.82 (0.78, 0.87) | **2.73E-12** |
| Phospholipids in Large HDL | 0.84 (0.80, 0.88) | **3.95E-11** |
| Phospholipids in Medium HDL | 1.01 (0.97, 1.05) | 6.69E-01 |
| Phospholipids in Small HDL | 1.09 (1.05, 1.14) | **2.59E-05** |
| **Total Lipids** |  |  |
| Total Lipids in Lipoprotein Particles | 1.07 (1.02, 1.11) | 2.74E-03 |
| Total Lipids in VLDL | 1.22 (1.17, 1.27) | **1.84E-20** |
| Total Lipids in Chylomicrons and Extremely Large VLDL | 1.26 (1.21, 1.30) | **2.47E-32** |
| Total Lipids in Very Large VLDL | 1.25 (1.20, 1.30) | **1.39E-29** |
| Total Lipids in Large VLDL | 1.22 (1.17, 1.27) | **1.27E-22** |
| Total Lipids in Medium VLDL | 1.12 (1.08, 1.17) | **3.87E-08** |
| Total Lipids in Small VLDL | 1.17 (1.12, 1.22) | **1.23E-13** |
| Total Lipids in Very Small VLDL | 1.10 (1.06, 1.15) | **8.50E-06** |
| Total Lipids in LDL | 0.98 (0.94, 1.03) | 4.15E-01 |
| Total Lipids in IDL | 0.94 (0.90, 0.98) | 6.73E-03 |
| Total Lipids in Large LDL | 0.96 (0.92, 1.00) | 5.84E-02 |
| Total Lipids in Medium LDL | 1.02 (0.98, 1.06) | 3.33E-01 |
| Total Lipids in Small LDL | 1.02 (0.98, 1.07) | 3.28E-01 |
| Total Lipids in HDL | 0.92 (0.88, 0.97) | 5.64E-04 |
| Total Lipids in Very Large HDL | 0.81 (0.77, 0.86) | **1.14E-13** |
| Total Lipids in Large HDL | 0.81 (0.77, 0.86) | **6.96E-15** |
| Total Lipids in Medium HDL | 0.97 (0.93, 1.02) | 2.20E-01 |
| Total Lipids in Small HDL | 1.07 (1.03, 1.12) | 5.19E-04 |
| **Sizes & apolipoprotein** |  |  |
| Average Diameter for VLDL Particles | 1.24 (1.19, 1.30) | **9.31E-23** |
| Average Diameter for LDL Particles | 0.85 (0.82, 0.88) | **1.64E-16** |
| Average Diameter for HDL Particles | 0.84 (0.80, 0.88) | **3.71E-11** |
| Apolipoprotein A1 | 0.94 (0.90, 0.99) | 1.03E-02 |
| Apolipoprotein B | 1.01 (0.97, 1.06) | 5.66E-01 |
| **Fatty acids** |  |  |
| Total Fatty Acids | 1.15 (1.10, 1.20) | **1.00E-11** |
| Omega-3 Fatty Acids | 1.02 (0.98, 1.07) | 2.63E-01 |
| Omega-6 Fatty Acids | 1.02 (0.98, 1.06) | 3.80E-01 |
| Polyunsaturated Fatty Acids | 1.02 (0.98, 1.07) | 3.07E-01 |
| Monounsaturated Fatty Acids | 1.20 (1.15, 1.25) | **1.58E-19** |
| Saturated Fatty Acids | 1.18 (1.14, 1.23) | **3.48E-17** |
| Linoleic Acid | 1.02 (0.98, 1.06) | 3.78E-01 |
| Docosahexaenoic Acid | 0.91 (0.87, 0.96) | **1.01E-04** |
| Degree of Unsaturation | 0.79 (0.75, 0.82) | **3.55E-28** |
| **Amino acids, cholines, and glycolysis** |  |  |
| Total Cholines | 1.02 (0.98, 1.06) | 3.79E-01 |
| Phosphatidylcholines | 1.04 (1.00, 1.09) | 6.02E-02 |
| Sphingomyelins | 0.89 (0.85, 0.94) | **1.15E-06** |
| Phosphoglycerides | 1.05 (1.01, 1.10) | 1.43E-02 |
| Glucose | 1.18 (1.13, 1.23) | **6.30E-16** |
| Lactate | 1.10 (1.06, 1.14) | **2.82E-06** |
| Pyruvate | 1.04 (1.00, 1.08) | 5.76E-02 |
| Citrate | 1.00 (0.96, 1.04) | 9.16E-01 |
| Alanine | 1.09 (1.05, 1.14) | **1.13E-05** |
| Glutamine | 0.93 (0.89, 0.96) | **1.42E-04** |
| Glycine | 0.84 (0.80, 0.89) | **6.37E-12** |
| Histidine | 0.99 (0.95, 1.03) | 5.47E-01 |
| Isoleucine | 1.07 (1.03, 1.12) | 3.25E-04 |
| Leucine | 1.09 (1.05, 1.13) | **1.75E-05** |
| Valine | 1.13 (1.09, 1.18) | **9.87E-10** |
| Phenylalanine | 1.03 (0.99, 1.07) | 1.67E-01 |
| Tyrosine | 1.12 (1.07, 1.16) | **3.59E-08** |
| Branched-Chain Amino Acids (Leucine + Isoleucine + Valine) | 1.11 (1.07, 1.16) | **1.36E-07** |
| **Ketone bodies & fluid balance** |  |  |
| 3-Hydroxybutyrate | 0.96 (0.92, 1.00) | 6.62E-02 |
| Acetate | 1.02 (0.98, 1.06) | 3.27E-01 |
| Acetoacetate | 0.99 (0.95, 1.03) | 4.70E-01 |
| Acetone | 0.94 (0.90, 0.98) | 4.28E-03 |
| Albumin | 1.07 (1.03, 1.12) | 7.61E-04 |
| Creatinine | 1.03 (0.99, 1.08) | 1.67E-01 |
| Glycoprotein Acetyls | 1.06 (1.01, 1.10) | 8.11E-03 |

Hazard ratios (HR) were presented per 1 standard deviation (SD) higher of metabolic biomarker on the natural log scale and were adjusted for age, sex, ethnicity, education, Townsend Deprivation Index, employment status, household income, family history of diabetes, history of CVD, history of hypertension, history of dyslipidemia, history of CLD, history of cancer, body mass index, waist circumference, hip circumference, smoking status, moderate alcohol, healthy diet score, healthy sleep score, physical activity, systolic blood pressure, diastolic blood pressure and glycated hemoglobin A1c. *P* value< 0.05/168 were highlighted in bold.

Apo-A1, apolipoprotein A1; Apo-B, apolipoprotein B; Apo-LP, apolipoprotein; BMI, body mass index; CVD, cardiovascular disease; CLD, chronic lung disease; DHA, docosahexaenoic acid; FA, fatty acids; HDL, high-density lipoproteins; HDL-D, high-density lipoprotein particle diameter; IDL, intermediate-density lipoproteins; L, large; LA, linoleic acid; LDL, low-density lipoproteins; LDL-D, low-density lipoprotein particle diameter; LP, lipoprotein; M, medium; MUFA, monounsaturated fatty acids; PUFA, polyunsaturated fatty acids; S, small; SFA, saturated fatty acids; VLDL, very-low-density lipoproteins; VLDL-D, very-low-density lipoprotein particle diameter; XL, very large; XS, very small; XXL, extremely large.
