## Supplementary file 3 for "Nuclear magnetic resonance-based metabolomics with machine learning for predicting progression from prediabetes to diabetes"

**Supplementary file 3. Coefficients of the selected 17 metabolites by priority-Lasso**.

| **Metabolites** | **Coefficient** |
| --- | --- |
| Average Diameter for LDL Particles | -0.031250025 |
| Sphingomyelins | -0.003620345 |
| Monounsaturated Fatty Acids | -0.073535676 |
| Docosahexaenoic Acid | -0.045470608 |
| Alanine | 0.004331897 |
| Glutamine | -0.034778028 |
| Glycine | -0.08627994 |
| Total Concentration of Branched-Chain Amino Acids (Leucine + Isoleucine + Valine) | -0.024140902 |
| Valine | -0.004270987 |
| Tyrosine | 0.035939862 |
| Glucose | 0.137870375 |
| Lactate | 0.105677477 |
| Triglycerides in Chylomicrons and Extremely Large VLDL | 0.022762656 |
| Triglycerides in Very Large VLDL | 0.002231071 |
| Cholesteryl Esters in Medium VLDL | -0.062412317 |
| Triglycerides in IDL | 0.193645548 |
| Cholesteryl Esters in Large HDL | -0.126588681 |

HDL, high-density lipoproteins; IDL, intermediate-density lipoproteins; LDL, low-density lipoproteins; VLDL, very-low-density lipoproteins.
