## Supplementary file 4 for "Nuclear magnetic resonance-based metabolomics with machine learning for predicting progression from prediabetes to diabetes"

| **Metabolites** | **HR (95% CI)** | ***P* value** |
| --- | --- | --- |
| Cholesteryl esters in large HDL | 0.72 (0.68, 0.76) | 6.54E-30 |
| Cholesteryl esters in medium VLDL | 0.86 (0.82, 0.90) | 3.71E-13 |
| Triglycerides in very large VLDL | 1.24 (1.20, 1.29) | 1.48E-29 |
| Average diameter for LDL particles | 0.84 (0.81, 0.87) | 5.77E-19 |
| Triglycerides in IDL | 1.19 (1.15, 1.24) | 4.03E-21 |
| Glycine | 0.84 (0.80, 0.88) | 9.74E-13 |
| Tyrosine | 1.11 (1.07, 1.15) | 2.35E-07 |
| Glucose | 1.16 (1.12, 1.21) | 6.07E-14 |
| Docosahexaenoic acid | 0.90 (0.86, 0.94) | 1.25E-06 |

**Supplementary file 4. Associations of the selected 9 metabolites with risk of diabetes among 13,489 participants with prediabetes after adjusting for conventional clinical variables.**

Hazard ratios (HR) were presented per 1 standard deviation (SD) higher of metabolic biomarker on the natural log scale and were adjusted for age, sex, Townsend Deprivation Index, family history of diabetes, body mass index, waist circumference, hip circumference, systolic blood pressure, diastolic blood pressure, and glycated hemoglobin A1c.

HDL, high-density lipoproteins; IDL, intermediate-density lipoproteins; LDL, low-density lipoproteins; VLDL, very-low-density lipoproteins.
