## Supplementary file 5 for "Nuclear magnetic resonance-based metabolomics with machine learning for predicting progression from prediabetes to diabetes"

| **Performance metric** | **Basic model^a^** | **Basic model + 9 metabolites^b^** | ***P* value** |
| --- | --- | --- | --- |
| AUROC |  |  |  |
| T=1-year | 0.821 (0.736, 0.907) | 0.868 (0.802, 0.934) | 0.016 |
| T=5-year | 0.790 (0.738, 0.842) | 0.811 (0.762, 0.860) | 0.033 |
| T=10-year | 0.791 (0.765, 0.816) | 0.806 (0.781, 0.831) | 0.006 |
| Continuous NRI |  |  |  |
| T=1-year | Reference | 0.266 (0.022, 0.599) | 0.036 |
| T=5-year | Reference | 0.217 (0.082, 0.348) | <0.001 |
| T=10-year | Reference | 0.199 (0.112, 0.274) | <0.001 |
| Absolute IDI |  |  |  |
| T=1-year | Reference | 0.002 (0.000, 0.005) | 0.052 |
| T=5-year | Reference | 0.003 (-0.003, 0.009) | 0.251 |
| T=10-year | Reference | 0.012 (0.006, 0.018) | <0.001 |

**Supplementary file 5. Performance of Cox proportional hazards prediction models for the risk of diabetes among participants with normoglycemia.**

^a^Basic model: age, sex, Townsend Deprivation Index, family history of diabetes mellitus, body mass index, Waist circumference, hip circumference, systolic blood pressure, diastolic blood pressure, and glycated hemoglobin A1c.

^b^The selected 9 metabolic biomarkers: cholesteryl esters in large HDL, triglycerides in very large VLDL, Glycine, average diameter for LDL particles, tyrosine, cholesteryl esters in medium VLDL, glucose, triglycerides in IDL, docosahexaenoic acid.

AUROC, area under the receiver operating characteristic curve; HDL, high-density lipoprotein; IDL, intermediate-density lipoprotein; IDI, absolute integrated discrimination improvement; LDL, low-density lipoprotein; NRI, net reclassification improvement; VLDL, very-low-density lipoprotein.
